# Use of tobacco and e-cigarettes among youth in Great Britain in 2022: analysis of a cross sectional survey

**DOI:** 10.1101/2022.07.25.22277745

**Authors:** Parris Williams, Hazel Cheeseman, Deborah Arnott, Laura Bunce, Nicholas S Hopkinson, Anthony A Laverty

**Affiliations:** National Heart and Lung Insistute, Imperial College London; Action on Smoking and Health; School of Public Health, Imperial College London

**Keywords:** E-cigarettes, Youth Tobacco use, Great Britain

## Abstract

**INTRODUCTION:** Although e-cigarettes can be an effective form of nicotine substitution for adults attempting to quit smoking, their use among children and young people is a concern. Accurate data about this are needed to inform debates over policy and regulation in the UK and elsewhere.

**METHODS:** Using data from an online survey of 2,613 people aged 11 to 18 years, conducted by the market research company *YouGov* in March 2022, we present prevalence estimates of e-cigarette and tobacco use.We use logistic regression models to assess differences in e-cigarette use, tobacco use and use of disposable e-cigarettes across a range of covariates including age, sex, tobacco smoking status, social class and country.

**RESULTS:** Among the 18.0% of those surveyed who reported ever having smoked a cigarette, 83.9% were not regular (at least once per week) smokers and 16.1% were (15.1% and 2.9% of the total sample respectively). Among the 19.2% of those surveyed who had ever used an e-cigarette, 79.2% were not regular users, while 20.8% were (15.2% and 4.0% respectively. Regular e-cigarette use was more common than regular tobacco smoking (4.0% vs 2.9%). E-cigarette use was more common among those who also smoked tobacco, with 9.0% of never e-cigarette users ever smoking tobacco, compared with 89.4% of regular e-cigarette users. Both smoking and e-cigarette use were associated with increasing age and use by others within the home, but not with social class. Use of disposable e-cigarettes was reported by 53.8% of those who have ever used an e-cigarette, and more common among girls than boys.

**DISCUSSION:** Regular e-cigarette use is now more common than smoking in children and youth, though the majority of this is among those who have also smoked tobacco. Measures to reduce the appeal of e-both e-cigarettes and tobacco to children and young people are warranted.

## INTRODUCTION

The use of e-cigarettes as a tool for harm reduction among people with tobacco dependence has gained recent momentum in the UK, and in 2021 the Medicines and Healthcare Products Regulatory agency (MHRA) announced that it would facilitate the process for e-cigarettes to become licensed as medicinal products, which would allow them to be marketed explicitly as smoking cessation aids and prescribed in smoking cessation clinics and elsewhere (1)

Although UK youth smoking rates are at an all time low (2), there are growing concerns that e-cigarette use may be developing among tobacco-naive young people, particularly the suggestion that e-cigarette use within this population may act as a gateway to cigarette smoking. In the US a 2018 report by the National Academies of Sciences, Engineering, and Medicine (NASEM) concluded that there was evidence for a causal effect of e-cigarette use on youth transition from non-smoker to ever smoker, while the Centers for Disease Control and Prevention (CDC) describe youth e-cigarette use as a serious public health concern(3, 4). Within the UK a 2020 PHE report stated that the majority of youth never smokers had also never used an e-cigarette with only 0.8%-1.3% of tobacco-naive youth reporting they were current e-cigarette users, concluding with a statement that e-cigarette prevalence among the UK’s young people needs to be closely monitored (5).

Given the importance of observing smoking and e-cigarette use within the younger population and the need for vigilance to track developments in the market, we report data from the 2022 ASH/YouGov Smokefree GB-Youth survey on youth smoking and e-cigarette use.

## METHODS

### Data

Data came from the ASH/YouGov Smokefree GB-Youth survey of 2,613 11-18 year olds, collected between 1^st^ and 29^th^ March 2022. Data were collected by YouGov, a public limited company, from an online panel recruited using proprietary software which is detailed elsewhere(6). Consent was provided by participants if they were aged over 16 years, and by their parents if 15 years or younger.

### Measures

E-cigarette use was assessed using the question *“Which ONE of the following is closest to describing your experience of e-cigarettes?”* We categorised this into never e-cigarette users (“I have never used an e-cigarette”), non-regular e-cigarette users (“I have only tried an e-cigarette once or twice” ; “I used e-cigarettes in the past but no longer do” ; “I use e-cigarettes more than once a month, but less than once a week” ; “I use e-cigarettes sometimes, but no more than once a month”), and regular use (“I use e-cigarettes more than once a week but not every day” ; “I use e-cigarettes every day”). We also classified respondents who replied that they had never heard of e-cigarettes as never e-cigarette users. Tobacco use was assessed using the same categories. We present regression analyses for both regular use and ever use of both tobacco and e-cigarettes.

Participants who reported ever use of e-cigarettes were also asked about the order of product use. We report the percentage of tobacco smoking among those choosing to select “I tried smoking a real cigarette before I first tried using an e-cigarette” compared with those who selected “I tried using an e-cigarette before I first tried smoking a real cigarette.”Ever e-cigarette users were also asked “Have you EVER used a disposable electronic cigarette (non-rechargeable)?” which was classified into yes and no.

Other data included were age group (11 to 13, 14 to 15, 16 to 17, and 18 years olds), sex, social class (based on the Registrar General classification of occupations and classified as ABC1 (higher) vs. C2DE (lower), and country of Great Britain (England, Scotland, Wales). We assessed household use of e-cigarettes and tobacco from the questions “Does anyone who currently lives in your home use an e-cigarette? (yes vs. no)” and “Who in your family, if anyone, smokes tobacco cigarettes at the moment?” For tobacco use in the household, we created a binary variable which categorised all of: “Mother (or female carer)”, “Father (or male carer)”, “Brother or sister”, “Grandparent” and “Other family member(s)” as yes. We also assessed exposure to e-cigarette promotion using the question “For the following question, by “being promoted”, we mean something that tries to increase interest in or demand for e-cigarettes.In which, if any, of the following places do you ever see e-cigarettes being promoted?” Participants could select more than one response and we categorised exposure as selecting at least one of Billboards; shops; Online; On buses; On TV; In newspapers/ magazines; or “Somewhere else”. We did not have data on exposure to tobacco advertising.

### Analysis

Descriptive statistics are presented after accounting for survey weighting to deal with differential non-response. Logistic regression analyses assessed differences in ever and regular e-cigarette and tobacco use and use of disposable e-cigarettes among those who reported ever use of e-cigarettes.

## RESULTS

The majority of participants lived in England (2281), with 206 in Scotland and 126 in Wales (Table 1). There were 1756 participants in ABC1 social class and 755 in C2DE. There were 341 participants aged 18 years old and the majority of participants (997) were aged 11 to 13 years.

**Table 1:** sample, numbers weighted using survey weighting.

|  | number | % |
| --- | --- | --- |
| Age |  |  |
| 11 to 13 years | 997 | 38.2 |
| 14 to 15 years | 625 | 23.9 |
| 16 to 17 years | 649 | 24.8 |
| 18 years | 341 | 13.1 |
| Sex |  |  |
| Male | 1340 | 51.3 |
| Female | 1273 | 48.7 |
| Social class |  |  |
| ABC1 (higher) | 1756 | 67.2 |
| C2DE (lower) | 755 | 28.9 |
| Country |  |  |
| England | 2281 | 87.3 |
| Scotland | 206 | 7.9 |
| Wales | 126 | 4.8 |
| Overall | 2613 | 100 |

Ever smoking tobacco was reported by 18.0% of the sample. 83.9% of these were non-regular smokers (absolute percentage 15.1%) and 16.1% were regular smokers (absolute percentage 2.9%) (Table 2). Among the 19.2% of those surveyed who had ever used an e-cigarette, 79.2% were not regular users, while 20.8% were (15.2% and 4.0% respectively). Regular e-cigarette use was more common than regular tobacco smoking (4.0% vs 2.9%). E-cigarette use was more common among those who also smoked tobacco, with 9.0% of never e-cigarette users ever smoking tobacco, compared with 89.4% of regular e-cigarette users.

**Table 2:** E-cigarette use (%) by tobacco smoking status.

|  | Never<br>smoked<br>tobacco | Non-regular | Regular<br>smoker | Ever<br>smoked<br>tobacco | Total |
| --- | --- | --- | --- | --- | --- |
| **Never used e-cigarette\*** | 91.0 | 8.5 | 0.5 | 9.0 | 80.9 |
| **Non regular e-cigarette user** | 24.4 | 57.9 | 17.8 | 75.7 | 15.2 |
| **Regular e-cigarette user** | 10.6 | 41.2 | 48.2 | 89.4 | 4.0 |
| **Ever e-cigarette user** | 35.4 | 52.6 | 12.0 | 64.6 | 19.2 |
| **Total** | 81.9 | 15.1 | 2.9 | 18.0 | 100 |

\*Never = "I have never used an e-cigarette", Non regular = "I have only tried an e-cigarette once or twice"; "I used e-cigarettes in the past but no longer do"; "I use e-cigarettes more

Questions on ordering of product use found that regular tobacco smoking was more common among those who reported using tobacco before e-cigarettes than those who used e-cigarettes first (20.2% vs 9.3%).

Differences in ever and regular use of e-cigarettes are shown in Table 3. Ever use was more common as participants got older (p for trend <0.001). Participants aged 18 years were considerably more likely to report ever use than those aged 11 to 13 years (adjusted odds ratio = 13.79, p<0.001). Ever use was more common among participants reporting regular e-cigarette use in their home (AOR = 6.65, p<0.001), although no differences were detected by social class or country. Associations were similar for regular use, with use more common among older participants and those reporting e-cigarette use inside the home (AOR = 20.05, p<0.001). Exposure to e-cigarette promotion was linked to greater likelihood of ever use of e-cigarettes (AOR = 1.61, p<0.001) with a weaker association with regular e-cigarette use (AOR = 1.52, p=0.097).

**Table 3:** Ever and regular use of e-cigarettes by selected characteristics.

|  | Ever use |  | Regular use |  |
| --- | --- | --- | --- | --- |
|  | Odds ratio | p value | Odds ratio | p value |
| Boys | ref | ref | ref | ref |
| Girls | 1.11 | 0.412 | 1.25 | 0.315 |
| 11 to 13 years | ref | ref | ref | ref |
| 14 to 15 years | 3.23 | <0.001 | 2.80 | 0.049 |
| 16 to 17 years | 7.93 | <0.001 | 9.54 | <0.001 |
| 18 years | 13.79 | <0.001 | 10.68 | <0.001 |
| England | ref | ref | ref | ref |
| Scotland | 1.23 | 0.279 | 1.02 | 0.949 |
| wales | 0.52 | 0.086 | 0.40 | 0.261 |
| Higher social class (ABC1) | ref | ref | ref | ref |
| Lower social class (C2DE) | 0.93 | 0.578 | 0.71 | 0.148 |
| Has not seen e-cigarettes promoted | ref | ref | ref | ref |
| Has seen e-cigarettes promoted* | 1.61 | <0.001 | 1.52 | 0.097 |
| No e-cigarette use in home | ref | ref | ref | ref |
| E-cigarette use in home | 6.65 | <0.001 | 20.75 | <0.001 |

\* In at least one of Billboards; shops; Online; On buses; On TV; In newspapers/ magazines; or “Somewhere else”

Differences in ever and regular tobacco smoking are shown in Table 4. These present a similar picture to that for e-cigarette use. Ever use was more common among older participants (e.g. AOR for 18 year olds vs. 11 to 13 year olds = 10.79, p<0.001) as well as those with cigarette smoking in the family (AOR = 2.42, p<0.001). Regular smoking was more common among older participants (p for trend <0.001) and those reporting cigarette smoking in the family (AOR = 3.87, p<0.001. We did not detect differences by sex or social class for either ever or regular tobacco smoking.

**Table 4:** Ever and regular tobacco smoking by selected characteristics.

|  | Ever smoking |  | Regular smoking |  |
| --- | --- | --- | --- | --- |
|  | Odds ratio | p value | Odds ratio | p value |
| Boys | ref | ref | ref | ref |
| Girls | 0.92 | 0.477 | 0.84 | 0.498 |
| 11 to 13 years | ref | ref | ref | ref |
| 14 to 15 years | 2.42 | <0.001 | 1.28 | 0.625 |
| 16 to 17 years | 6.50 | <0.001 | 3.80 | 0.002 |
| 18 years | 10.79 | <0.001 | 5.81 | <0.001 |
| England | ref | ref | ref | ref |
| Scotland | 1.18 | 0.349 | 1.33 | 0.414 |
| Wales | 0.51 | 0.042 | 0.61 | 0.405 |
| Higher social class (ABC1) | ref | ref | ref | ref |
| Lower social class (C2DE) | 1.01 | 0.966 | 1.11 | 0.677 |
| No tobacco cigarette smoking in family | ref | ref | ref | ref |
| Tobacco cigarette smoking in family | 2.42 | <0.001 | 3.87 | <0.001 |

Differences in use of disposable e-cigarettes among ever e-cigarette users are shown in Table 5. Use of disposable products was more common among girls (AOR = 1.69, p=0.010), 18 year olds (AOR = 2.78, p = 0.020), and those reporting e-cigarette use in the home (AOR = 2.60, p<0.001).

**Table 5:** use of disposable e-cigarettes among ever e-cigarette users.

|  | % | Odds ratio | p value |
| --- | --- | --- | --- |
| Boys | 50.9 | ref | ref |
| Girls | 59.3 | 1.69 | 0.010 |
| 11 to 13 years | 41.1 | ref | ref |
| 14 to 15 years | 45.7 | 1.29 | 0.581 |
| 16 to 17 years | 59.6 | 2.33 | 0.052 |
| 18 years | 62.5 | 2.78 | 0.020 |
| England | 54.9 | ref | ref |
| Scotland | 56.7 | 0.80 | 0.433 |
| Wales | 60.3 | 1.79 | 0.457 |
| Higher social class (ABC1) | 53.3 | ref | ref |
| Lower social class (C2DE) | 58.8 | 1.24 | 0.313 |
| Has not seen e-cigarettes promoted | 54.8 | ref | ref |
| Has seen e-cigarettes promoted | 56.6 | 1.22 | 0.400 |
| No e-cigarette use in home | 45.6 | ref | Ref |
| E-cigarette use in home | 66.3 | 2.60 | <0.001 |

\* In at least one of Billboards; shops; Online; On buses; On TV; In newspapers/ magazines; or "Somewhere else"

## DISCUSSION

Youth tobacco smoking and e-cigarette usage remained low in 2022, however e-cigarette usage in this population is now slightly higher than regular tobacco smoking. Compared to data from 2020-21 the low smoking prevalence is unchanged and regular e-cigarette use is also similar (4.0% vs 4.3%,(7). Most e-cigarette users report that they have experimented with or are regular tobacco users alongside their e-cigarette use.

9.3% of those who reported that they used e-cigarettes before smoking tobacco were also regular tobacco smokers. Despite this low percentage it is still a cause for concern and has been previously demonstrated across the West. A 2017 US meta-analysis concluded that adolescents who reported any vaping experience (both regular and intermittent), increased their odds of subsequent cigarette smoking by a third (8). Data from our survey demonstrates an increase in young people trying e-cigarettes first, in 2018 18.3% of youth reported they tried e-cigarettes before smoking (7), this has now increased to 37.5%. Despite youth smoking rates being at an all time low, these increases in e-cigarette use without previous tobacco smoking are a potential cause for concern. While it may be the case that there is common susceptibility to both tobacco and e-cigarette use, there is a need for regulation and enforcement to ensure lower levels of e-cigarette use among children. Current regulation in the UK prohibits the sale of nicotine containing e-cigarettes to those under the age of 18 (9). While we did not assess whether e-cigarettes used by study respondents contained nicotine, previous analyses of this data found that 76% of participants reported that their e-cigarettes always or sometimes contained nicotine.(10) These high levels of use strongly suggest that current laws are poorly enforced and underscore recent recommendations for increased investment in local smokefree enforcement actions (11).

Age is also a significant factor regarding e-cigarette and smoking behaviour. As demonstrated in previous (7) and by our current survey data, 18-year-olds are significantly more likely to report ever smoking and e-cigarette use compared with lower age groups. We found that use of e-cigarettes by others within the home was associated with e-cigarette use among youth, and the same was true for tobacco smoking. This is not surprising and is a well-known risk factor for smoking behaviour in young people, observed in both the UK and Germany (12, 13). We also found that reporting exposure to promotion of e-cigarettes was linked to ever use of these products. This finding underscores the need to strengthen enforcement of the rules on advertising, promotion and enforcement, and review whether they need further strengthening.

Disposable e-cigarette use in the US and UK is growing, recent data from a cross sectional survey of adults in Great Britain concluded that disposable e-cigarette use grew significantly in 2022, predominantly in 18 year olds (14). We found that, among ever e-cigarette users, use of disposables was reported by more than half of respondents, more commonly among girls and 18 year olds. More research is needed to understand the reasons and risk factors for disposable e-cigarette use among younger populations.

Limitations of the current study include the survey study design meaning that we cannot infer causality. This means that we are unable to determine trajectories of tobacco smoking, e-cigarette use and quitting of products.It is plausible that some e-cigarette use in this population is among tobacco smokers attempting to quit, or may replace trying smoking, and further research into these issues is needed. However, the study sample was designed to be representative of the Great Britain population and these results add to the ongoing surveillance of smoking and e-cigarette use among the UK’s younger population, which is vital for understanding and informing tobacco control policy.

In conclusion, rates of e-cigarette use among young people are now higher than smoking, though the vast majority of regular e-cigarette users also smoke or have done in the past. Measures are needed to reduce the appeal of these devices to young people as well as enforcement of measures to prevent illegal sales and marketing.

## Data Availability

All data produced in the present study are available upon reasonable request to the authors

---

**Figure 1:**
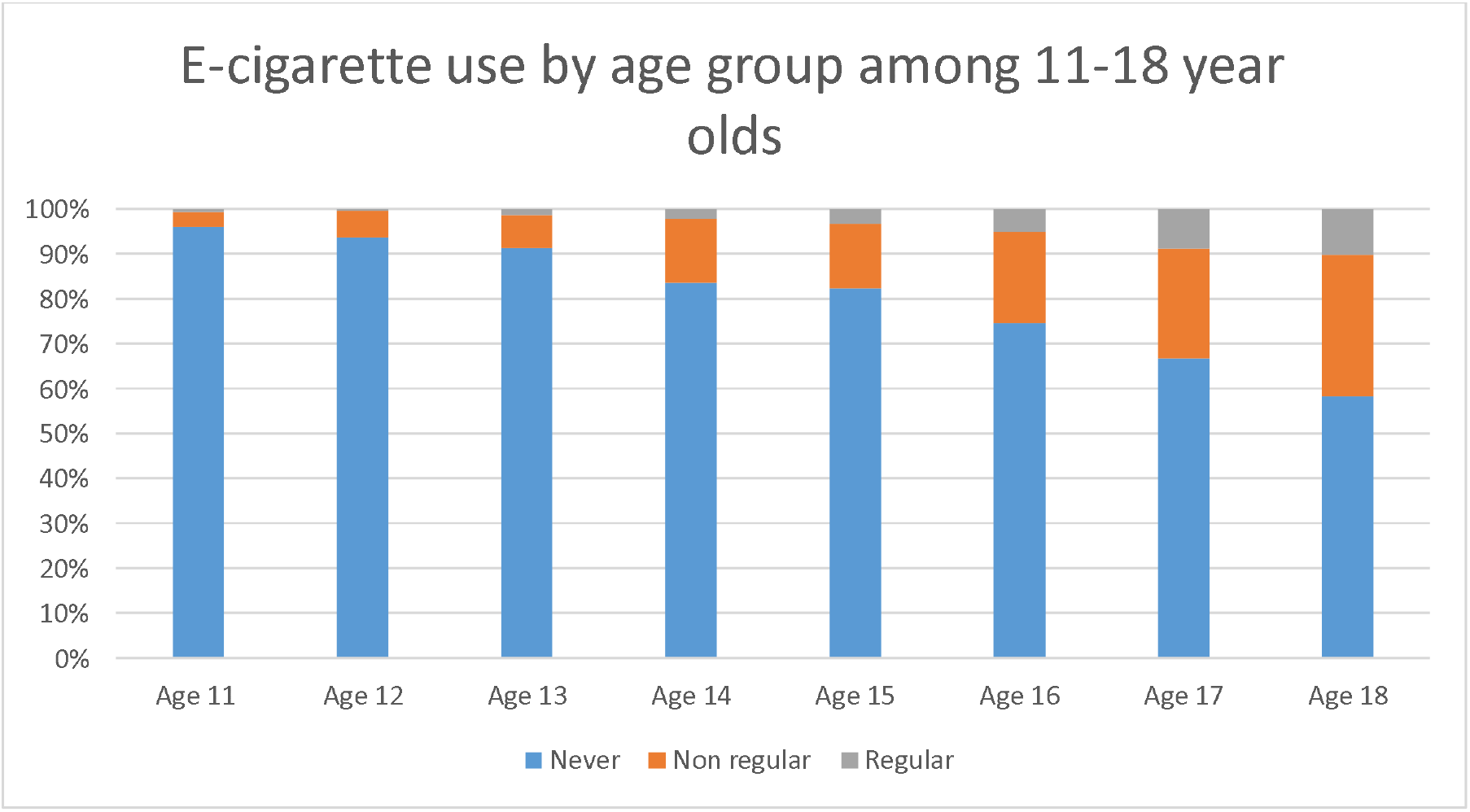
E-cigarette use by age. *Never = “I have never used an e-cigarette”, Non regular = “I have only tried an e-cigarette once or twice”; “I used e-cigarettes in the past but no longer do” ; “I use e-cigarettes more than once a month, but less than once a week” ; “I use e-cigarettes sometimes, but no more than once a month”, Regular = “I use e-cigarettes more than once a week but not every day” ; “I use e-cigarettes every day”. Respondents who responded that they had never heard of e-cigarettes also classified as never e-cigarette users.

## Notes

### Competing Interest Statement

NSH is Chair of Action on Smoking and Health and Medical Director of The British Lung Foundation. AAL is a Trustee of Action on Smoking and Health. Other authors have no conflict of interest to declare

### Funding Statement

This study did not receive any funding

### Author Declarations

Ethics committee of Imperial College Research Governance and Integrity Team name gave ethical approval for this work (ICREC Ref: 20IC6625)

## References

1. MHRA publishes clear guidance to support bringing e-cigarettes to market as licensed therapies [press release]. Gov.UK 2021.

2. NHS Digital. Smoking patterns among young people 2019 [Available from: https://digital.nhs.uk/data-and-information/publications/statistical/statistics-on-smoking/statistics-on-smoking-england-2019/part-4-smoking-patterns-in-children-copy#:~:text=Smoking%20prevalence%20among%20young%20peopl,-Smoking%20prevalence%2C%20by&text=6%25%20of%20pupils%20were%20current,3%25%20were%20regular2%20smokers.&text=Similar%20proportions%20of%20boys%20and,7%25%20of%2015%20year%20olds.

3. National Academies of Sciences E, Medicine. Public health consequences of e-cigarettes. 2018.

4. Centers for disease control. Youth E-Cigarette Use Remains Serious Public Health Concern Amid COVID-19 Pandemic 2021 [Available from: https://www.cdc.gov/media/releases/2021/p0930-e-cigarette.html#:~:text=A%20study%20released%20today%20(attached,those%20youth%20using%20flavored%20e%2D.

5. Public Health England. Vaping in England: 2021 evidence update summary 2021 [Available from: https://www.gov.uk/government/publications/vaping-in-england-evidence-update-february-2021/vaping-in-england-2021-evidence-update-summary.

6. Eastwood B, Dockrell M, Arnott D, Britton J, Cheeseman H, Jarvis MJ, et al. Electronic cigarette use in young people in Great Britain 2013–2014. Public Health. 2015;129(9):1150–6.

7. Action on Smoking and Health. Use of e-cigarettes among young people in Great Britain 2021 [Available from: https://ash.org.uk/wp-content/uploads/2021/07/Use-of-e-cigarettes-among-young-people-in-Great-Britain-2021.pdf.

8. Soneji S, Barrington-Trimis JL, Wills TA, Leventhal AM, Unger JB, Gibson LA, et al. Association between initial use of e-cigarettes and subsequent cigarette smoking among adolescents and young adults: a systematic review and meta-analysis. JAMA pediatrics. 2017;171(8):788–97.

9. British Goverment. THE NICOTINE INHALING PRODUCTS (AGE OF SALE AND PROXY PURCHASING) REGULATIONS 2015 In: Care DoHaS, editor. 2015.

10. Action on Smoking and Health. Use of e-cigarettes (vapes) among young people in Great Britain 2022 [Available from: https://ash.org.uk/wp-content/uploads/2022/07/Use-of-e-cigarettes-among-young-people-in-Great-Britain-2022.pdf.

11. Disparities OfHIa. The Khan review, Making smoking obsolete In: Disparities OfHIa, editor. Gov.UK 2022.

12. Hanewinkel R, Isensee B. Risk factors for e-cigarette, conventional cigarette, and dual use in German adolescents: a cohort study. Preventive medicine. 2015;74:59–62.

13. Laverty AA, Filippidis FT, Taylor-Robinson D, Millett C, Bush A, Hopkinson NS. Smoking uptake in UK children: analysis of the UK Millennium Cohort Study. Thorax. 2019;74(6):607–10.

14. Tattan-Birch H, Jackson SE, Kock L, Dockrell M, Brown J. Rapid growth in disposable e-cigarette vaping among young adults in Great Britain from 2021 to 2022: a repeat cross-sectional survey. medRxiv. 2022.

